# Clinical Features and Outcomes of Neuroleptospirosis in French Guiana: Insights from a Retrospective Study (2015-2021)

**DOI:** 10.64898/2026.04.24.26350928

**Authors:** Audrey Delahaye, Tanguy Dequidt, Fabrice Quet, Clementine Montagnac, Theo Blaise, Hatem Kallel, Felix Djossou, Mathieu Nacher, Kinda Schepers, Julien Coussement, Paul Le Turnier, Loic Epelboin

**Author notes:** Corresponding author (ADE) Audrey Delahaye, Department of Infectious and Tropical Diseases, CHU de Guyane. 3 avenue Alexis Blaise, Cayenne 97300, French Guiana, France.

## Abstract

**Background:** Leptospirosis is a re-emerging zoonosis in French Guiana, with broad and sometimes misleading clinical spectrum. Neurological involvement, referred to as neuroleptospirosis, lacks a consensus definition. Therefore, it is likely underrecognized, raising concerns about missed diagnosis and potentially poor outcomes. This study aimed to characterize the epidemiology, clinical features and outcomes of neuroleptospirosis.

**Methodology/ Principal findings:** A multicenter retrospective study was conducted across all public healthcare facilities of French Guiana (2015-2021). Neuroleptospirosis was defined by the combination of (i) neurological symptoms and (ii) objective evidence of neurological involvement based on cerebrospinal fluid (CSF) analysis and/or neuroimaging. Cases were compared with leptospirosis without neurological involvement. Among 146 consecutive hospital-managed cases of leptospirosis, 18 (12%) met criteria for neuroleptospirosis (incidence 0.88 cases/100,000 inhabitants/year; 95% CI 0.52-1.39). Among them, 78% (14/18) presented a meningeal syndrome, 22% (4/18) an encephalitic syndrome, and 17% (3/18) showed paresthesia. Lumbar puncture was performed in 28/146 (19%) patients, with pleocytosis observed in 18/28 (64%) patients; median CSF leukocyte count was 42 cells/mm^3^ (range 13-240/mm^3^), with lymphocytic predominance in 8/13 (62%), a slight protein level (median 0.51 g/l, range 0.32-0.88) and no hypoglycorachia. Brain MRI revealed abnormalities in 2/8 (25%) cases of neuroleptospirosis: one cerebral infarct and one pachymeningitis. Compared with patients without neurological involvement, factors associated with a diagnosis of neuroleptospirosis were an age <30 years (*p=*0.004, 95% CI 1.72-18), no leukocytosis (*p=*0.042, 95% CI 1.04-10.39) nor thrombocytopenia (*p=*0.012, 95% CI 1.39-14.32) during hospitalization. Neuroleptospirosis cases had milder disease: they less often progressed to sepsis (OR 0.28; 95% CI 0.10-0.79); none required intensive care admission nor died.

**Discussion/Conclusions:** This study provides new insights into the clinical spectrum and outcomes of neuroleptospirosis in French Guiana. Prospective studies incorporating a consensus definition, systematic CSF analysis and microbiological testing are warranted to further characterize the pathophysiology and optimize diagnostic strategies for neuroleptospirosis.

**Author summary:** Leptospirosis is a re-emerging infection in French Guiana, with a broad range of clinical presentations. Yet, its neurological manifestations—referred to as neuroleptospirosis — remain likely underrecognized, lacking a consensus definition. We conducted a multicenter retrospective study to describe the epidemiological, clinical features and outcomes of neuroleptospirosis. Then, we compared cases with non-neurological leptospirosis. We used a standardized definition of neuroleptospirosis, based on neurological symptoms associated with cerebrospinal fluid analysis and/or neuroimaging. Neuroleptospirosis accounted for one tenth of leptospirosis cases. Clinical presentation was largely dominated by meningitis (three quarters of cases). Laboratory parameters showed lower rates of leukocytosis and renal involvement. CSF profiles revealed moderate lymphocytic pleocytosis, closely mimicking viral infections. Strikingly, compared with non-neurological forms, neuroleptospirosis cases exhibited fewer severity markers and uniformly favorable outcomes, with lower sepsis, no deaths nor need for intensive care admission. Factors associated with a diagnosis of neuroleptospirosis were an age lower than 30, the absence of hyperleukocytosis and thrombocytopenia during hospitalization. Neuroleptospirosis accounted for a non-negligible proportion of leptospirosis cases in French Guiana, exhibiting CSF features mimicking viral meningitis, and carrying excellent outcomes. Increasing awareness and improving recognition of this presentation is essential to reduce underdiagnosis and refine patient management in endemic settings.

## Introduction

Leptospirosis is a globally distributed bacterial zoonosis caused by pathogenic spirochaetes of the genus *Leptospira* [1]. In French Guiana, the disease is endemic and its incidence has markedly increased in recent years, from 2.4/100,000 in 2016 to 10.4/100,000 in 2022 [2–4]. The clinical presentation of leptospirosis is highly heterogeneous and nonspecific, ranging from mild, self-limited illness to life-threatening disease [1]. While classical clinical forms are well described, atypical manifestations may lead to delayed or misdiagnosis.

Neurological involvement, referred to as neuroleptospirosis, remains poorly recognized and is likely underdiagnosed among central nervous system (CNS) infections. In patients presenting primarily with neurological symptoms associated with fever, bacterial or viral meningitis are often considered the primary diagnosis. As leptospires are very sensitive to antibiotic treatment and *Leptospira* Polymerase Chain Reaction (PCR) is not included in the CSF multiplex panel, the diagnosis of leptospirosis can be missed, even if efficiently treated by an antibiotic treatment given for suspicion of classic bacterial meningitis [5,6]. Therefore, neuroleptospirosis may be overlooked [7–10]. Differential diagnosis presents challenges in tropical regions, where multiple infections can affect the CNS, including arboviruses and malaria [11]. In South America, several studies conducted in Brazil have retrospectively reported leptospirosis as a common cause of aseptic meningitis (clinical signs and symptoms of meningitis with pleocytosis but negative Gram stain and sterile bacterial cultures) [12–14].

A wide spectrum of neurological complications has been associated with leptospirosis, including meningitis, encephalitis, stroke, Guillain-Barre syndrome, acute disseminated encephalomyelitis, myelitis or cranial nerve palsies, and uveitis [8,10,15–21]. Neuroleptospirosis is especially prone to under or misdiagnosis when neurological manifestations are isolated, without concomitant systemic features suggestive of leptospirosis [7,8]. Delayed recognition may limit timely initiation of appropriate therapy and adversely affect outcomes [10]. Improved characterization of the clinical and laboratory features of neuroleptospirosis is therefore essential to enhance physician awareness and diagnostic accuracy.

The primary objective of this study was to characterize the epidemiology, clinical features and outcomes of neuroleptospirosis. The secondary objective was to explore the factors associated with neurological involvement in leptospirosis.

## Methods

### Ethics statement

The study was registered with the Health Data Hub (No. F20220426211656). All patients were informed about their right to object the use of their data. This retrospective study relied exclusively on routinely collected healthcare data and involved no intervention or additional biological sample collection. The study complied with the French reference methodology MR-004 for research using health data not involving direct participation and was declared to the French Data Protection Authority (Commission Nationale de l’ Informatique et des Libertes, CNIL) under the above registration number. The data used in this study were de-identified prior to their use, and all procedures complied with applicable ethical standards.

### Study design

This multicenter study was conducted across all the public healthcare facilities of the territory, i.e. the three public hospitals of French Guiana (Cayenne, Kourou, and Saint-Laurent-du-Maroni), including the 17 hospital-affiliated remote prevention and care centers (Centres Delocalises de Prevention et de Soins, CDPS), as well as the three community hospitals located across most of the inland municipalities, between 2015 and 2021. All consecutive hospital-managed leptospirosis cases meeting inclusion criteria during the study period were included. Ambulatory patients managed exclusively in primary care settings (i.e., general practitioners) were not included. Exclusion criteria were age under 18 years at diagnosis, patient refusal to participate, or incomplete medical records.

### Inclusion criteria

In accordance with World Health Organization (WHO) and Centers for Disease Control and Prevention (CDC)’ guidelines, the diagnosis of leptospirosis was based on the presence of compatible clinical features, in the absence of a more likely alternate diagnosis, combined with microbiological evidence [22,23]. Microbiological evidence of leptospirosis was defined as either a positive PCR (in a serum, urine, or CSF), and/or a positive anti-*Leptospira* immunoglobulin M (IgM) serology, with or without confirmation by microscopic agglutination test (MAT)[4,24].

Leptospirosis cases were divided as either confirmed or probable. Confirmed leptospirosis was defined by either a positive PCR (serum, urine, and/or CSF) or by serological criteria combining an IgM level ≥ 50 IU/mL or IgM seroconversion with a MAT titer ≥ 1:200, a MAT seroconversion ≥ 1:400, or at least a fourfold increase in MAT titer. Alternatively, a confirmed case could be defined by two IgM ELISA tests ≥ 100 IU/mL taken at least seven days apart. Probable leptospirosis was defined, in patients presenting with compatible clinical symptoms and no alternative diagnosis, by an IgM level ≥ 50 IU/mL or IgM seroconversion with either MAT ≥ 1:100 (and < 1:200), MAT seroconversion ≥ 1:100 (and < 1:400), or a fourfold increase in MAT titer. Alternatively, a probable case could be defined by a single IgM ≥ 100 IU/mL if MAT is unavailable.

Neuroleptospirosis was defined by (i) a confirmed or probable diagnosis of leptospirosis associated with (ii) neurological signs and (iii) abnormal CSF findings and/or brain or spinal imaging abnormalities attributable to leptospirosis. Neurological syndromes were classified according to the guidelines of the French Infectious Diseases Society (SPILF) [25]. A meningeal syndrome was defined by the presence of at least two of the following symptoms: headache, neck stiffness, or vomiting. An encephalitic syndrome was defined by seizures, impaired consciousness, or focal neurological deficits; patients fulfilling both definitions were classified as having meningoencephalitis. CSF abnormalities were defined using standard thresholds: pleocytosis as a white cell count ≥ 5 cells/mm^3^, elevated protein concentration as ≥ 0.45 g/L and hypoglycorrhachia as CSF glucose < 40% of concomitant blood glucose levels. Additional CSF parameters were interpreted according to local laboratory reference ranges, including lactate (1.1-2.4 mmol/L) and IgG (10-30 mg/L). Brain and spinal imaging abnormalities were defined according to the interpretation of the attending radiologist.

Severe leptospirosis was defined as need for vasopressors or mechanical ventilation, initiation of renal replacement therapy, or outcome of death [2]. The Sequential Organ Failure Assessment (SOFA) score was retrospectively calculated when data were available, with sepsis defined as a SOFA score ≥ 2 [26]. Acute Respiratory Distress Syndrome (ARDS) was defined according to Berlin criteria [27]. Antibiotic therapy considered active against leptospirosis included β-lactams (excluding aztreonam), tetracyclines, macrolides, fluoroquinolones, and aminoglycosides [28].

### Microbiological analyses

Biological samples were analyzed according to the usual standard care procedure for the region at Cerba Laboratory and Biomnis Laboratory (mainland France). Both laboratories performed in-house-real-time PCR assays targeting the *Leptospira* 16S rRNA and *lipL32* genes, as well as anti-*Leptospira* IgM serology using an enzyme-linked immunosorbent assay (ELISA) (Serion, Germany). ELISA results were interpreted as positive for titers ≥ 50 IU/mL, intermediate for titers between 15 and 50 IU/mL, and negative for titers ≤ 15 IU/mL [29,30]. MAT was performed either at Eurofins Biomnis or at the French National Reference Center for Leptospirosis (Institut Pasteur, Paris), using a standard serogroup panel. The full list of tested serogroups is provided in **Appendix 1**. The presumptive infecting serogroup was defined by the highest MAT titer detected in the most recent serum sample; identical titers across multiple serogroups were classified as coagglutinins.

### Data collection and study procedures

Patients were identified using ICD-10 coding data from participating healthcare centers, cross-referenced with laboratory results. Subsequently, electronic medical records were manually screened for eligibility. For included patients, demographic, exposure, clinical, laboratory, imaging, management and outcome data were retrospectively collected using a standardized, pseudo-anonymized case report form. Baseline clinical and non-clinical findings were obtained within the first 48 hours of the initial healthcare encounter, and extreme (maximum sand minimums) laboratory values during hospitalization were also documented.

### Statistical analysis

Quantitative variables were summarized as medians and interquartile ranges, and qualitative variables as frequencies and percentages with *n* indicating the number of available observations for each variable. No imputation was performed for missing data. Comparisons of quantitative variables were made using Student’s t-test for normally distributed variables or the Mann-Whitney *U* test for non-normal distributions or sample sizes <40. Qualitative variables were compared using the chi-squared test or Fisher’s exact test. All tests were two-tailed, and p-values ≤ 0.05 were considered statistically significant. Statistical analyses were performed using Stata version 19 (StataCorp LLC, College Station, TX, USA). An exploratory multivariable analysis was performed using logistic regression to identify factors associated with neuroleptospirosis. A backward stepwise approach was applied, initially including all variables with p ≤ 0.20 in bivariate analyses, while excluding redundant variables. Continuous variables, including laboratory parameters, were categorized using clinically relevant thresholds reported in the literature or, when available, according to the median or mean value depending on their distribution. Variable selection was guided by the Akaike information criterion (AIC), retaining variables whose removal resulted in an increase in the AIC. Model fit was assessed using the Hosmer-Lemeshow goodness-of-fit test. Incidence rates of neuroleptospirosis per 100,000 inhabitants per year were estimated using population data from the National Institute of Statistics and Economic Studies (INSEE).

## Results

Over the 7-year study period, 146 consecutive hospital-managed cases of leptospirosis were identified, among which 18 patients (12%) met the study criteria for neuroleptospirosis (**Figure 1**). The incidence of hospital-managed neuroleptospirosis was 0.88 per 100,000 inhabitants per year (95% CI 0.52-1.39).

**Figure 1.**
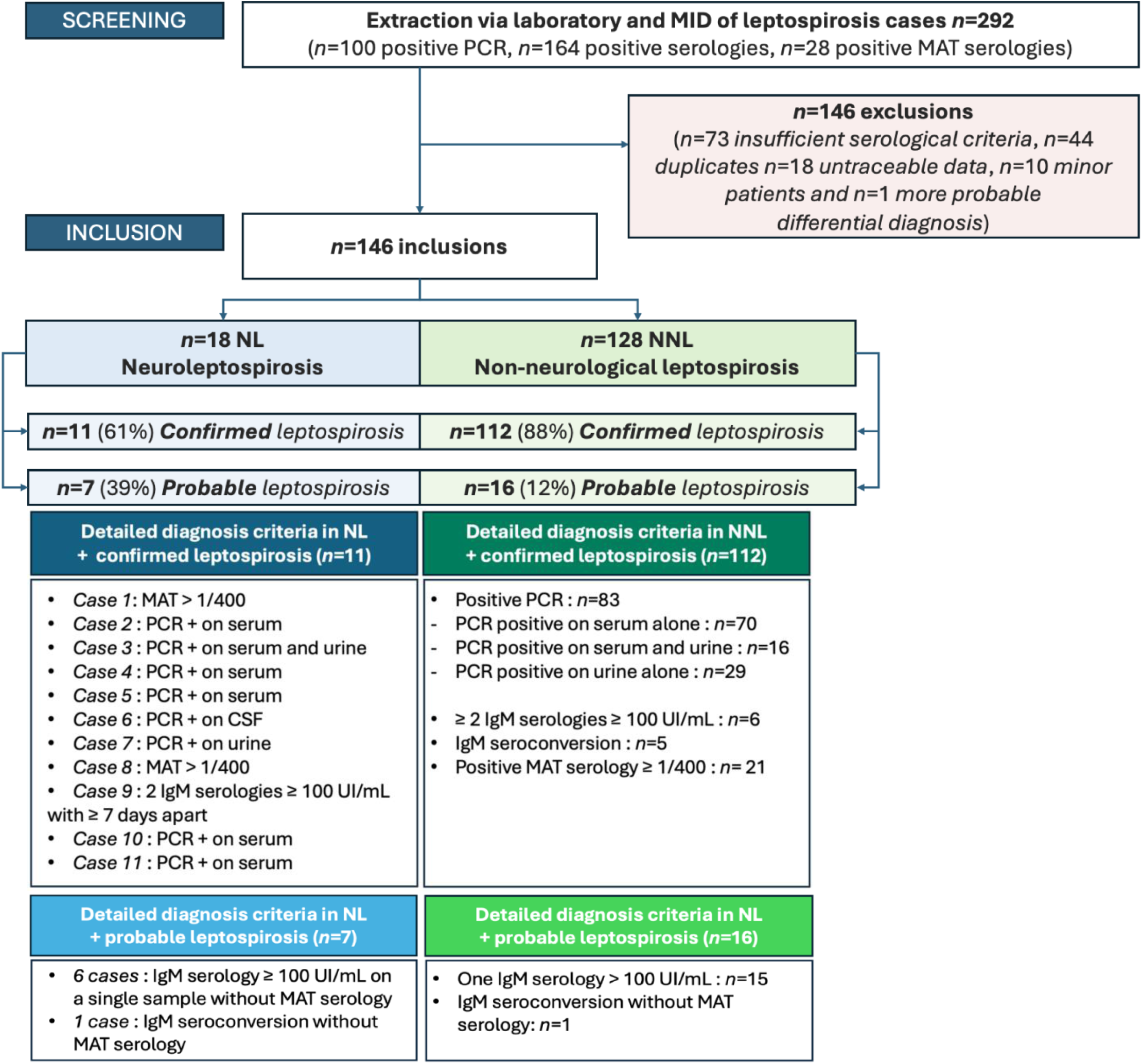
Flowchart. *n/N* corresponds to the number of patients. **CSF**, cerebrospinal fluid, **MID**, medical information department - **IgM**, immunoglobulins M - **MAT**, microscopic agglutination test - **NL**, neuroleptospirosis case - **NNL**, non-neuroleptospirosis case - **PCR**, polymerase chain reaction. **Duplicates**: patients identified simultaneously by the laboratory and through extraction from the MID. **Neuroleptospirosis case (NL):** presence of (i) confirmed or probable leptospirosis (as defined above), associated with (ii) neurological signs, and abnormal CSF findings and/or brain or spinal imaging abnormalities attributable to leptospirosis. **Confirmed leptospirosis:** positive PCR (serum, urine, and/or CSF); IgM level ≥ 50 IU/mL; or IgM seroconversion*, each of the latter two criteria requiring either a MAT titer ≥ 1:200, a MAT seroconversion** ≥ 1:400, or a fourfold increase in MAT titer. Alternatively, a confirmed case could be defined by two IgM tests ≥ 100 IU/mL taken at least seven days apart. **Probable leptospirosis:** IgM level ≥ 50 IU/mL or IgM seroconversion* with either MAT ≥ 1:100 (and < 1:200), MAT seroconversion ≥ 1:100 (and < 1:400), or a fourfold increase in MAT titer. Alternatively, a probable case could be defined by a single IgM ≥ 100 IU/mL if MAT is unavailable. * **IgM seroconversion:** negative or equivocal IgM level (15-50 IU/mL) on the first sample, followed by a positive result (≥ 50 IU/mL) on the second sample collected at least seven days later. ** **MAT seroconversion:** initial MAT < 1:100 on the first sample, becoming positive on a second sample collected at least seven days later.

Epidemiological and exposure characteristics of patients with neuroleptospirosis are described in **Table 1**. Patients with neuroleptospirosis had a median age of 26 years (interquartile range IQR: 23-35). Male-to-female ratio was 5. Half of patients were born in Brazil (8/16, 50%), and 25% in French Guiana (4/16). Most patients had an at-risk exposure: occupational (80%), environmental (67%), or animal (80%).

**Table 1.**
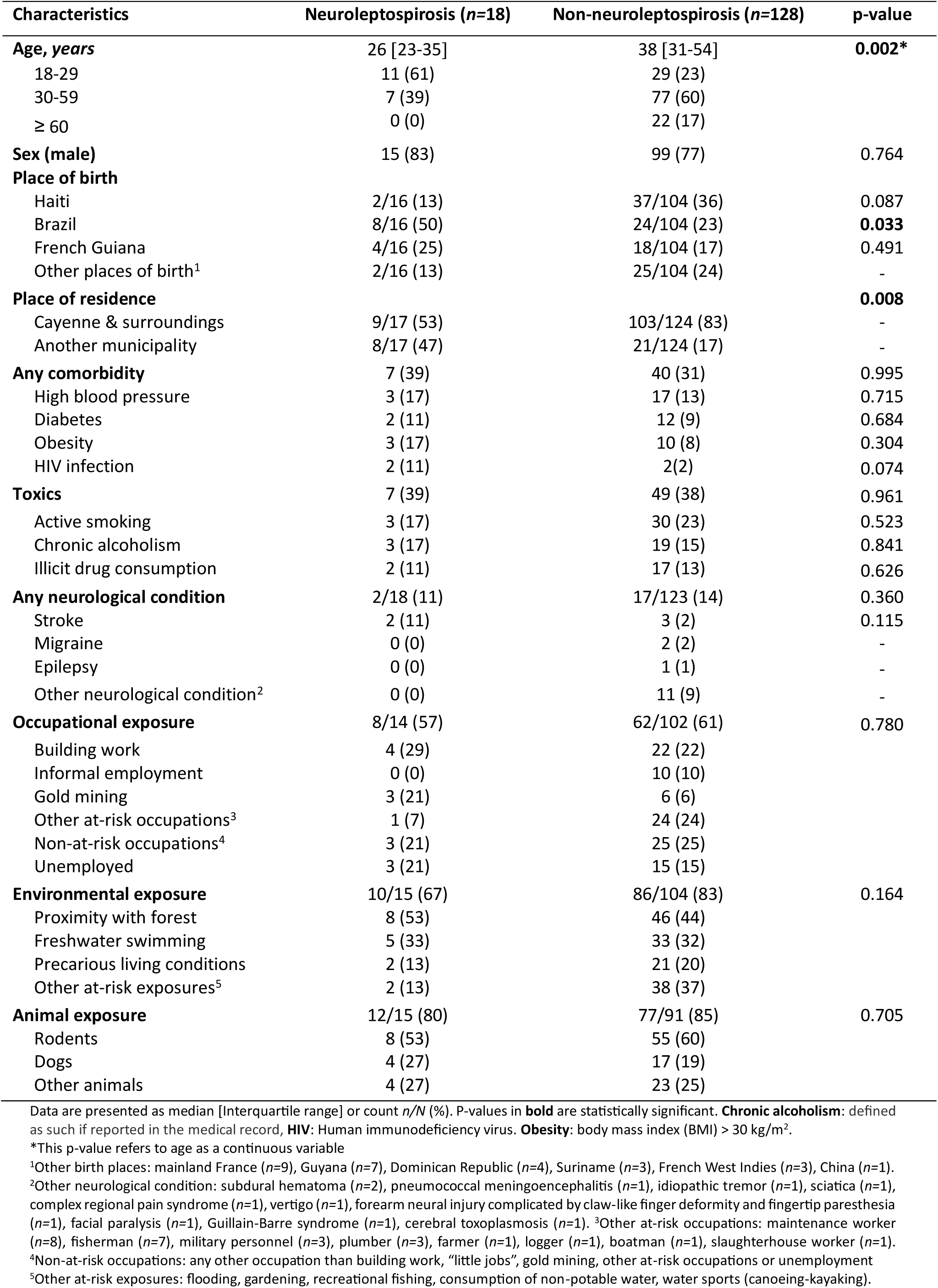
Socio-demographic and exposure characteristics.

| Characteristics | Neuroleptospirosis (n=18) | Non-neuroleptospirosis (n=128) | p-value |
| --- | --- | --- | --- |
| **Age, years** | 26 [23-35] | 38 [31-54] | **0.002\*** |
| 18-29 | 11 (61) | 29 (23) |  |
| 30-59 | 7 (39) | 77 (60) |  |
| ≥ 60 | 0 (0) | 22 (17) |  |
| **Sex (male)** | 15 (83) | 99 (77) | 0.764 |
| **Place of birth** |  |  |  |
| Haiti | 2/16 (13) | 37/104 (36) | 0.087 |
| Brazil | 8/16 (50) | 24/104 (23) | **0.033** |
| French Guiana | 4/16 (25) | 18/104 (17) | 0.491 |
| Other places of birth<sup>1</sup> | 2/16 (13) | 25/104 (24) | - |
| **Place of residence** |  |  | **0.008** |
| Cayenne & surroundings | 9/17 (53) | 103/124 (83) | - |
| Another municipality | 8/17 (47) | 21/124 (17) | - |
| **Any comorbidity** | 7 (39) | 40 (31) | 0.995 |
| High blood pressure | 3 (17) | 17 (13) | 0.715 |
| Diabetes | 2 (11) | 12 (9) | 0.684 |
| Obesity | 3 (17) | 10 (8) | 0.304 |
| HIV infection | 2 (11) | 2(2) | 0.074 |
| **Toxics** | 7 (39) | 49 (38) | 0.961 |
| Active smoking | 3 (17) | 30 (23) | 0.523 |
| Chronic alcoholism | 3 (17) | 19 (15) | 0.841 |
| Illicit drug consumption | 2 (11) | 17 (13) | 0.626 |
| **Any neurological condition** | 2/18 (11) | 17/123 (14) | 0.360 |
| Stroke | 2 (11) | 3 (2) | 0.115 |
| Migraine | 0 (0) | 2 (2) | - |
| Epilepsy | 0 (0) | 1 (1) | - |
| Other neurological condition<sup>2</sup> | 0 (0) | 11 (9) | - |
| **Occupational exposure** | 8/14 (57) | 62/102 (61) | 0.780 |
| Building work | 4 (29) | 22 (22) |  |
| Informal employment | 0 (0) | 10 (10) |  |
| Gold mining | 3 (21) | 6 (6) |  |
| Other at-risk occupations<sup>3</sup> | 1 (7) | 24 (24) |  |
| Non-at-risk occupations<sup>4</sup> | 3 (21) | 25 (25) |  |
| Unemployed | 3 (21) | 15 (15) |  |
| **Environmental exposure** | 10/15 (67) | 86/104 (83) | 0.164 |
| Proximity with forest | 8 (53) | 46 (44) |  |
| Freshwater swimming | 5 (33) | 33 (32) |  |
| Precarious living conditions | 2 (13) | 21 (20) |  |
| Other at-risk exposures<sup>5</sup> | 2 (13) | 38 (37) |  |
| **Animal exposure** | 12/15 (80) | 77/91 (85) | 0.705 |
| Rodents | 8 (53) | 55 (60) |  |
| Dogs | 4 (27) | 17 (19) |  |
| Other animals | 4 (27) | 23 (25) |  |

Data are presented as median [Interquartile range] or count n/N (%). P-values in **bold** are statistically significant. **Chronic alcoholism**: defined as such if reported in the medical record, **HIV**: Human immunodeficiency virus. **Obesity**: body mass index (BMI) > 30 kg/m<sup>2</sup>.
\*This p-value refers to age as a continuous variable
<sup>1</sup>Other birth places: mainland France (n=9), Guyana (n=7), Dominican Republic (n=4), Suriname (n=3), French West Indies (n=3), China (n=1).
<sup>2</sup>Other neurological condition: subdural hematoma (n=2), pneumococcal meningoencephalitis (n=1), idiopathic tremor (n=1), sciatica (n=1), complex regional pain syndrome (n=1), vertigo (n=1), forearm neural injury complicated by claw-like finger deformity and fingertip paresthesia (n=1), facial paralysis (n=1), Guillain-Barre syndrome (n=1), cerebral toxoplasmosis (n=1). <sup>3</sup>Other at-risk occupations: maintenance worker (n=8), fisherman (n=7), military personnel (n=3), plumber (n=3), farmer (n=1), logger (n=1), boatman (n=1), slaughterhouse worker (n=1).
<sup>4</sup>Non-at-risk occupations: any other occupation than building work, "little jobs", gold mining, other at-risk occupations or unemployment
<sup>5</sup>Other at-risk exposures: flooding, gardening, recreational fishing, consumption of non-potable water, water sports (canoeing-kayaking).

Clinical manifestations are described in **Tables 2 & 3.** In patients with neuroleptospirosis, neurological symptoms were generally present on the first day of illness (*n=*16, 89%), and always during the first week. Meningeal syndrome was the most frequent clinical presentation (*n=*14, 78%). Encephalitic syndrome occurred in a quarter of patients (*n=*4, 22%), including two with impaired consciousness (minimum Glasgow coma score - GCS - of 9) and three with focal deficits: one patient developed peripheral facial diplegia with dysarthria, swallowing, and gait disturbances, another presented acute right hemiplegia with left central facial paralysis consistent with an ischemic stroke; and one patient had a transient unilateral sudden visual loss. Peripheral nervous system symptoms were observed alongside CNS involvement in three patients (17%), consisting of distal paresthesia.

Laboratory parameters showed that patients with neuroleptospirosis did not exhibit hyperleukocytosis (median 8.1 x 10^9^/L, IQR 5.7-11.3) nor thrombocytopenia (median 200 x 10^9^/L, IQR 70-227). Only mild hepatic or renal abnormalities were present (**Table 2**). CSF analysis revealed pleocytosis in most patients (16/18, 89%) with a median of 42 cells/mm³ (IQR 13-240). Overall, CSF profiles were predominantly lymphocytic (8/13, 62%). More specifically, during the first week of illness, 4/7 (57%) of CSF samples were neutrophilic, 2/7 (29%) lymphocytic and 1/7 (14%) mixed. By contrast, from day 8 onward, all samples were lymphocytic, 6/6 (100%) (**Appendix 2 & Table 3**). In addition, CSF analysis showed a mild protein level elevation (median 0.51 g/L, IQR 0.32-0.88), and a normal glucose level. The limited number of MAT serologies tests - due to a loss of reimbursement for the technique in 2014 - precluded assessment of potential neurotropic strains (**Appendix 4**).

**Table 2.**
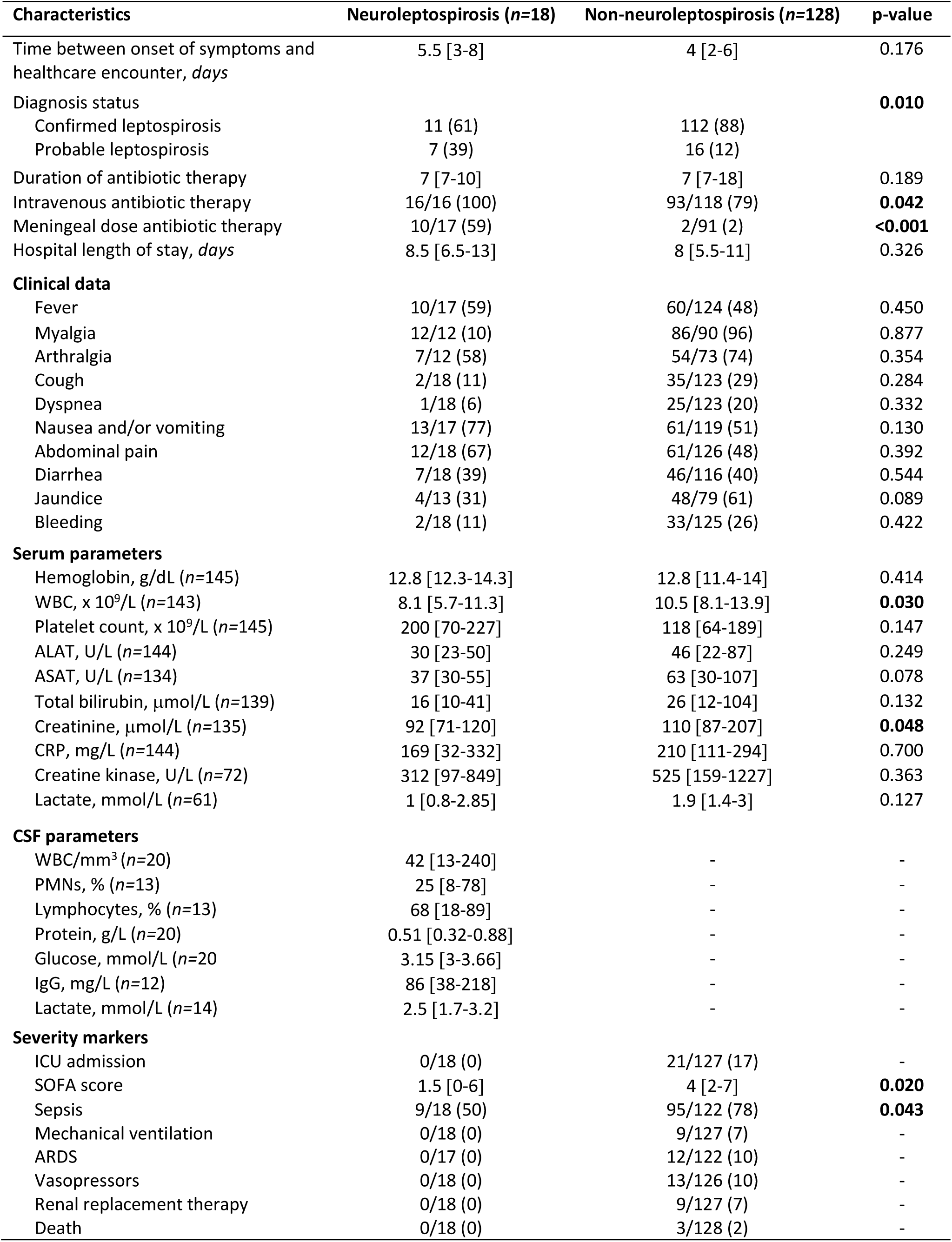
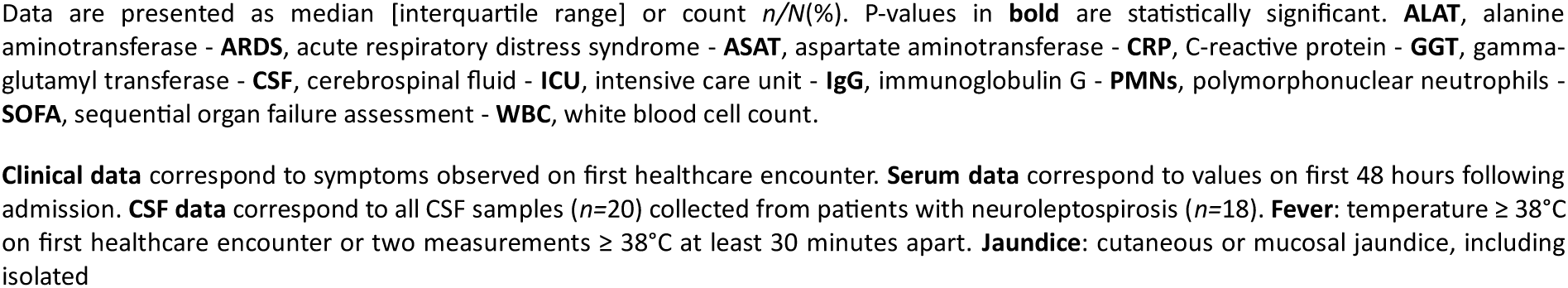
Clinical signs, laboratory and severity parameters.

| Characteristics | Neuroleptospirosis (n=18) | Non-neuroleptospirosis (n=128) | p-value |
| --- | --- | --- | --- |
| Time between onset of symptoms and healthcare encounter, *days* | 5.5 [3-8] | 4 [2-6] | 0.176 |
| Diagnosis status |  |  | **0.010** |
| Confirmed leptospirosis | 11 (61) | 112 (88) |  |
| Probable leptospirosis | 7 (39) | 16 (12) |  |
| Duration of antibiotic therapy | 7 [7-10] | 7 [7-18] | 0.189 |
| Intravenous antibiotic therapy | 16/16 (100) | 93/118 (79) | **0.042** |
| Meningeal dose antibiotic therapy | 10/17 (59) | 2/91 (2) | **<0.001** |
| Hospital length of stay, *days* | 8.5 [6.5-13] | 8 [5.5-11] | 0.326 |
| **Clinical data** |  |  |  |
| Fever | 10/17 (59) | 60/124 (48) | 0.450 |
| Myalgia | 12/12 (10) | 86/90 (96) | 0.877 |
| Arthralgia | 7/12 (58) | 54/73 (74) | 0.354 |
| Cough | 2/18 (11) | 35/123 (29) | 0.284 |
| Dyspnea | 1/18 (6) | 25/123 (20) | 0.332 |
| Nausea and/or vomiting | 13/17 (77) | 61/119 (51) | 0.130 |
| Abdominal pain | 12/18 (67) | 61/126 (48) | 0.392 |
| Diarrhea | 7/18 (39) | 46/116 (40) | 0.544 |
| Jaundice | 4/13 (31) | 48/79 (61) | 0.089 |
| Bleeding | 2/18 (11) | 33/125 (26) | 0.422 |
| **Serum parameters** |  |  |  |
| Hemoglobin, g/dL (n=145) | 12.8 [12.3-14.3] | 12.8 [11.4-14] | 0.414 |
| WBC, x 10<sup>9</sup>/L (n=143) | 8.1 [5.7-11.3] | 10.5 [8.1-13.9] | **0.030** |
| Platelet count, x 10<sup>9</sup>/L (n=145) | 200 [70-227] | 118 [64-189] | 0.147 |
| ALAT, U/L (n=144) | 30 [23-50] | 46 [22-87] | 0.249 |
| ASAT, U/L (n=134) | 37 [30-55] | 63 [30-107] | 0.078 |
| Total bilirubin, µmol/L (n=139) | 16 [10-41] | 26 [12-104] | 0.132 |
| Creatinine, µmol/L (n=135) | 92 [71-120] | 110 [87-207] | **0.048** |
| CRP, mg/L (n=144) | 169 [32-332] | 210 [111-294] | 0.700 |
| Creatine kinase, U/L (n=72) | 312 [97-849] | 525 [159-1227] | 0.363 |
| Lactate, mmol/L (n=61) | 1 [0.8-2.85] | 1.9 [1.4-3] | 0.127 |
| **CSF parameters** |  |  |  |
| WBC/mm<sup>3</sup> (n=20) | 42 [13-240] | - | - |
| PMNs, % (n=13) | 25 [8-78] | - | - |
| Lymphocytes, % (n=13) | 68 [18-89] | - | - |
| Protein, g/L (n=20) | 0.51 [0.32-0.88] | - | - |
| Glucose, mmol/L (n=20) | 3.15 [3-3.66] | - | - |
| IgG, mg/L (n=12) | 86 [38-218] | - | - |
| Lactate, mmol/L (n=14) | 2.5 [1.7-3.2] | - | - |
| **Severity markers** |  |  |  |
| ICU admission | 0/18 (0) | 21/127 (17) | - |
| SOFA score | 1.5 [0-6] | 4 [2-7] | **0.020** |
| Sepsis | 9/18 (50) | 95/122 (78) | **0.043** |
| Mechanical ventilation | 0/18 (0) | 9/127 (7) | - |
| ARDS | 0/17 (0) | 12/122 (10) | - |
| Vasopressors | 0/18 (0) | 13/126 (10) | - |
| Renal replacement therapy | 0/18 (0) | 9/127 (7) | - |
| Death | 0/18 (0) | 3/128 (2) | - |

Data are presented as median [interquartile range] or count $n/N(\%)$ . P-values in **bold** are statistically significant. **ALAT**, alanine aminotransferase - **ARDS**, acute respiratory distress syndrome - **ASAT**, aspartate aminotransferase - **CRP**, C-reactive protein - **GGT**, gamma- glutamyl transferase - **CSF**, cerebrospinal fluid - **ICU**, intensive care unit - **IgG**, immunoglobulin G - **PMNs**, polymorphonuclear neutrophils - **SOFA**, sequential organ failure assessment - **WBC**, white blood cell count.
**Clinical data** correspond to symptoms observed on first healthcare encounter. **Serum data** correspond to values on first 48 hours following admission. **CSF data** correspond to all CSF samples ( $n=20$ ) collected from patients with neuroleptospirosis ( $n=18$ ). **Fever**: temperature $\geq 38^{\circ}\text{C}$ on first healthcare encounter or two measurements $\geq 38^{\circ}\text{C}$ at least 30 minutes apart. **Jaundice**: cutaneous or mucosal jaundice, including isolated

**Table 3.**
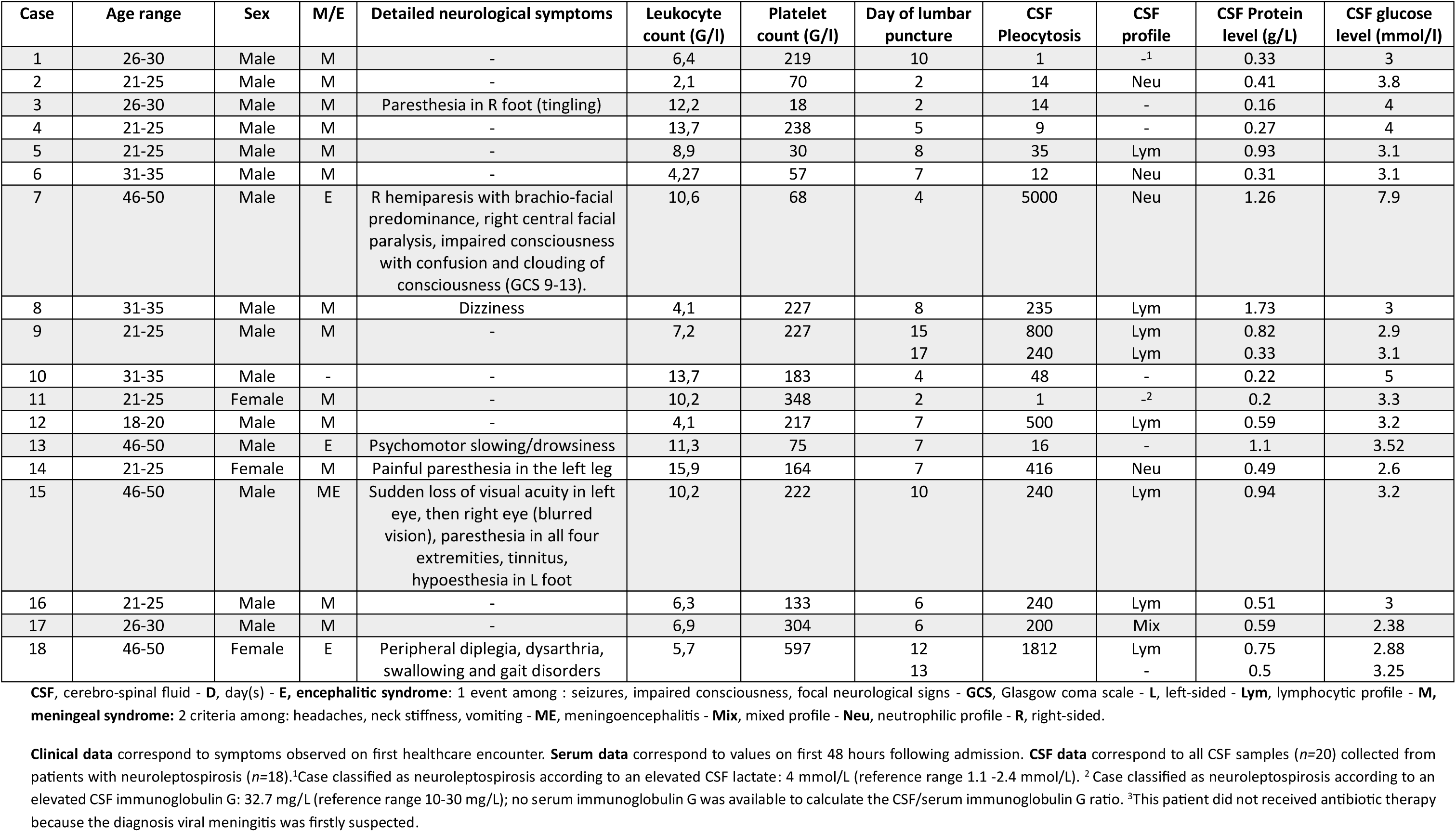
Clinical and biological characteristics (serum and CSF) of the 18 cases of neuroleptospirosis.

| Case | Age range | Sex | M/E | Detailed neurological symptoms | Leukocyte count (G/l) | Platelet count (G/l) | Day of lumbar puncture | CSF Pleocytosis | CSF profile | CSF Protein level (g/L) | CSF glucose level (mmol/l) |
| --- | --- | --- | --- | --- | --- | --- | --- | --- | --- | --- | --- |
| 1 | 26-30 | Male | M | - | 6,4 | 219 | 10 | 1 | -<sup>1</sup> | 0.33 | 3 |
| 2 | 21-25 | Male | M | - | 2,1 | 70 | 2 | 14 | Neu | 0.41 | 3.8 |
| 3 | 26-30 | Male | M | Paresthesia in R foot (tingling) | 12,2 | 18 | 2 | 14 | - | 0.16 | 4 |
| 4 | 21-25 | Male | M | - | 13,7 | 238 | 5 | 9 | - | 0.27 | 4 |
| 5 | 21-25 | Male | M | - | 8,9 | 30 | 8 | 35 | Lym | 0.93 | 3.1 |
| 6 | 31-35 | Male | M | - | 4,27 | 57 | 7 | 12 | Neu | 0.31 | 3.1 |
| 7 | 46-50 | Male | E | R hemiparesis with brachio-facial predominance, right central facial paralysis, impaired consciousness with confusion and clouding of consciousness (GCS 9-13). | 10,6 | 68 | 4 | 5000 | Neu | 1.26 | 7.9 |
| 8 | 31-35 | Male | M | Dizziness | 4,1 | 227 | 8 | 235 | Lym | 1.73 | 3 |
| 9 | 21-25 | Male | M | - | 7,2 | 227 | 15<br>17 | 800<br>240 | Lym<br>Lym | 0.82<br>0.33 | 2.9<br>3.1 |
| 10 | 31-35 | Male | - | - | 13,7 | 183 | 4 | 48 | - | 0.22 | 5 |
| 11 | 21-25 | Female | M | - | 10,2 | 348 | 2 | 1 | -<sup>2</sup> | 0.2 | 3.3 |
| 12 | 18-20 | Male | M | - | 4,1 | 217 | 7 | 500 | Lym | 0.59 | 3.2 |
| 13 | 46-50 | Male | E | Psychomotor slowing/drowsiness | 11,3 | 75 | 7 | 16 | - | 1.1 | 3.52 |
| 14 | 21-25 | Female | M | Painful paresthesia in the left leg | 15,9 | 164 | 7 | 416 | Neu | 0.49 | 2.6 |
| 15 | 46-50 | Male | ME | Sudden loss of visual acuity in left eye, then right eye (blurred vision), paresthesia in all four extremities, tinnitus, hypoesthesia in L foot | 10,2 | 222 | 10 | 240 | Lym | 0.94 | 3.2 |
| 16 | 21-25 | Male | M | - | 6,3 | 133 | 6 | 240 | Lym | 0.51 | 3 |
| 17 | 26-30 | Male | M | - | 6,9 | 304 | 6 | 200 | Mix | 0.59 | 2.38 |
| 18 | 46-50 | Female | E | Peripheral diplegia, dysarthria, swallowing and gait disorders | 5,7 | 597 | 12<br>13 | 1812 | Lym<br>- | 0.75<br>0.5 | 2.88<br>3.25 |

CSF, cerebro-spinal fluid - **D**, day(s) - **E**, **encephalitic syndrome**: 1 event among : seizures, impaired consciousness, focal neurological signs - **GCS**, Glasgow coma scale - **L**, left-sided - **Lym**, lymphocytic profile - **M**, **meningeal syndrome**: 2 criteria among: headaches, neck stiffness, vomiting - **ME**, meningoencephalitis - **Mix**, mixed profile - **Neu**, neutrophilic profile - **R**, right-sided.
**Clinical data** correspond to symptoms observed on first healthcare encounter. **Serum data** correspond to values on first 48 hours following admission. **CSF data** correspond to all CSF samples ( $n=20$ ) collected from patients with neuroleptospirosis ( $n=18$ ).<sup>1</sup>Case classified as neuroleptospirosis according to an elevated CSF lactate: 4 mmol/L (reference range 1.1 -2.4 mmol/L). <sup>2</sup> Case classified as neuroleptospirosis according to an elevated CSF immunoglobulin G: 32.7 mg/L (reference range 10-30 mg/L); no serum immunoglobulin G was available to calculate the CSF/serum immunoglobulin G ratio. <sup>3</sup>This patient did not received antibiotic therapy because the diagnosis viral meningitis was firstly suspected.

Radiology parameters of patients with neuroleptospirosis (**Appendix 3)** were available from brain imaging, including CT-Scan and MRI (11 patients, 61%) and spinal MRI imaging (2 patients, 11%). All brain CT-scans (*n=*11) were normal. Of the five MRIs performed additionally, two revealed acute abnormalities. One patient with peripheral facial diplegia showed diffuse pachymeningitis with bilateral facial nerves enhancement on MRI (**Figure 2**). Another patient with stroke had multiple recent ischemic lesions in the left superficial middle cerebral artery territory, with a proximal thrombus. Regarding treatment, the median duration of antibiotic therapy was 7 days [IQR 7–10], frequently at meningeal doses (10/15, 67%). Most frequently used antibiotics were third-generation cephalosporins (14/17, 82%), penicillins (8/17, 47%), and doxycycline (3/17, 18%).

**Figure 2.**
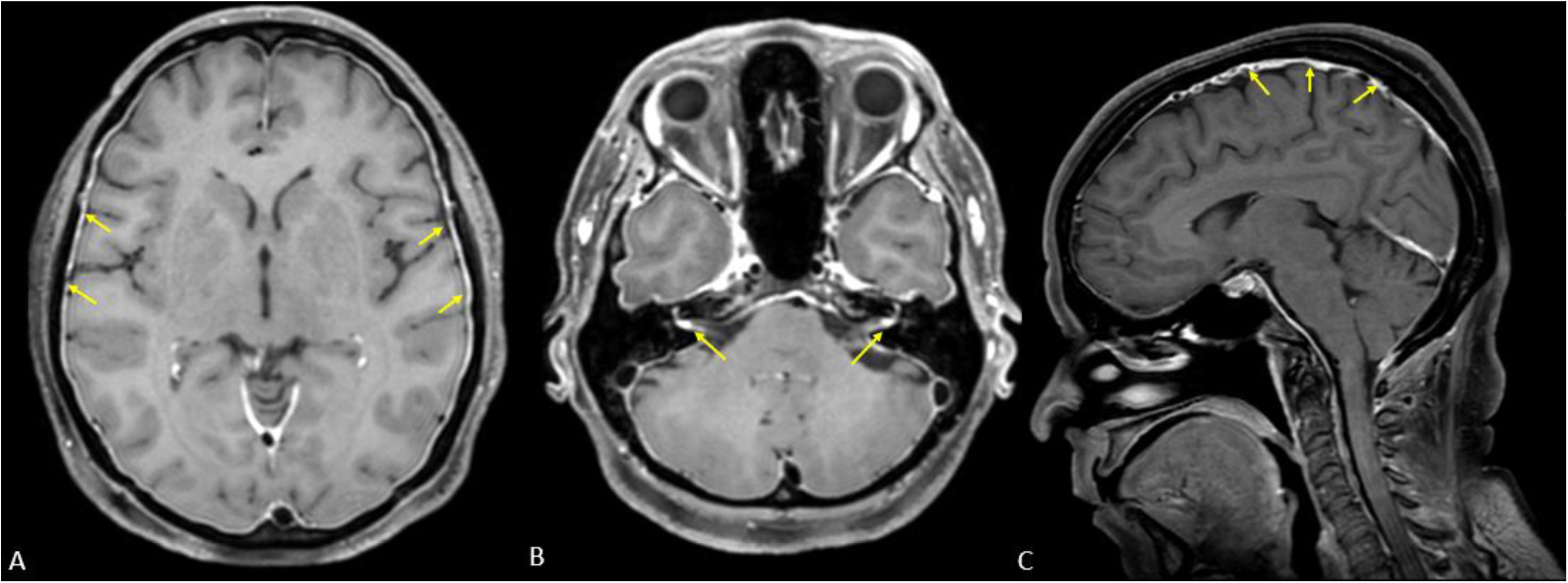
MRI images showing the pachymeningitis and the enhancement of facial nerves (Image analysis by Dr Rasouly Najibullah) **A.** Axial T1-weighted turbo spin echo (TSE) sequence showing thickening and inflammatory enhancement of the pachymeningeal layer. **B.** Axial T1 TSE sequence showing bilateral enhancement of the facial nerves. **C.** Sagittal T1 TSE sequence showing thickening and inflammatory enhancement of the pachymeninges.

Regarding the outcome of neuroleptospirosis (**Table 2 & Appendix 3**), none of the cases required ICU admission nor died during hospital stay. One patient developed persistent facial diplegia. This patient was lost to follow-up and had showed no improvement after one month of hospitalization. Among treated patients, 10/17 (59%) received meningeal-dose therapy and recovered without sequelae; 6/17 (35%) recovered despite receiving non-meningeal doses of antibiotic therapy. One patient, who was initially suspected to have viral meningitis, recovered without antibiotic treatment.

Patients with and without a diagnosis of neuroleptospirosis were compared.

Patients with neuroleptospirosis were younger (median age 26 vs 38, p=0.002), their white blood cell counts and median creatinine levels were significantly lower (8.1 vs 10.5 x 10 9 /L and 92 vs 110 μmol/L, respectively), sepsis was less frequent and SOFA scores were lower (p=0.043 vs. p=0.020) than in patients with non-neuroleptospirosis. (**Tables 1 & 2**). Multivariate analysis identified neuroleptospirosis to be independently associated with age under 30 years, absence of hyperleukocytosis, and absence of thrombocytopenia (**Table 4**).

**Table 4.** Factors evaluated for a diagnosis of neuroleptospirosis (bivariate and multivariable analysis)

| Variable | Bivariate analysis |  |  | Multivariate analysis |  |  | 338 |
| --- | --- | --- | --- | --- | --- | --- | --- |
|  | Crude ORs | 95% CI | p-value | Adjusted ORs | 95% CI | p-value | 339 |
| Age < 30 years | 5.36 | [1.90-15.08] | **0.001\*** | 5.56 | [1.72-18] | **0.004** | 340 |
| Confirmed diagnosis | 0.22 | [0.08-0.66] | **0.007\*** | - | - | - | 341 |
| Dyspnea | 0.23 | [0.03-1.82] | 0.164 | - | - | - | 342 |
| Bleeding | 0.35 | [0.08-1.60] | 0.175 | - | - | - | 343 |
| Nausea-vomiting | 3.09 | [0.95-10.03] | 0.060 | - | - | - | 344 |
| SOFA Score | 0.83 | [0.70-0.99] | **0.036\*** | - | - | - | 345 |
| Absence of leukocytosis | 4.29 | [1.49-12.32] | **0.007\*** | 3.29 | [1.04-10.39] | **0.042** | 346 |
| Absence of thrombocytopenia | 3.59 | [1.29-9.94] | **0.014\*** | 4.46 | [1.39-14.32] | **0.012** | 347 |
| ASAT< 60 U/L (2N) | 2.41 | [0.85-6.83] | 0.097 | - | - | - |  |
| ALAT< 70 U/L (2N) | 2.24 | [0.70-7.2] | 0.175 | - | - | - |  |
| Absence of jaundice | 2.48 | [0.82-7.5] | 0.107 | - | - | - |  |
| Absence of acute renal failure | 4.15 | [1.37-12.63] | **0.012\*** | - | - | - |  |

**Crude** odds ratios correspond to the bivariate model, while **adjusted** odds ratios correspond to the multivariate model. P-values in **bold** are statistically significant.
**ALAT**, alanine aminotransferase - **ASAT**, aspartate aminotransferase - **CI**, confidence interval - **NL**, neuroleptospirosis case - **NNL**, non-neuroleptospirosis case - **ORs**, odds ratio - **SOFA score**, Sequential Organ Failure Assessment score assessing six organ systems (respiratory, coagulation, liver, cardiovascular, central nervous system, and renal), each scored from 0 to 4, with higher scores indicating greater organ dysfunction. - **2N**, two times the normal value.
**Absence of leukocytosis**: maximum white blood cell count recorded during hospitalization < 10 x 10<sup>9</sup>/L - **Absence of thrombocytopenia**: minimum platelet count recorded during hospitalization > 150 x 10<sup>9</sup>/L. **Absence** **of jaundice**: maximum total bilirubin level recorded during hospitalization < 40 µmol/L - **Absence of ARF** : maximum creatinine level recorded during hospitalization < 130 µmol/L - **Neuroleptospirosis**: presence of (i) confirmed or probable leptospirosis (as defined above), (ii) clinical signs or symptoms consistent with neurological involvement, and abnormal CSF findings and/or brain or spinal imaging abnormalities attributable to leptospirosis.

## Discussion

This study provides a comprehensive description of the epidemiology, clinical presentation, and outcomes of neuroleptospirosis in an endemic setting.

Using stringent composite inclusion criteria combining clinical and CSF criteria, neurological involvement accounted for approximately one tenth of hospital-managed leptospirosis cases. This estimate is consistent with previous reports, which generally range from 5% to 20% of hospitalized leptospirosis cases [8,10,31]. The exclusion of ambulatory cases may have resulted in a biased estimate of this proportion. Nevertheless, studies indicate that most leptospirosis cases in French Guiana are treated as inpatients (73-86%) [2,4]. Comparison of our findings with those from previous studies is limited by the absence of a consensus to define neuroleptospirosis, with some studies relying exclusively on clinical manifestations, and others incorporating CSF criteria [8,31].

We hypothesize that the high proportion of patients born in Brazil and gold miners (mostly originating from Brazil) in the neuroleptospirosis group may reflect an increased exposure to sylvatic strains, which may be more neurotropic, rather than genetic susceptibility, [32,33].

In our series, neurological manifestations presented most often early in the clinical course, thereby challenging the classical biphasic model with temporal separation between bacteremic and immune phases, considering the neurologic component of leptospirosis as a delayed immune complication [1,34]. Meningeal presentations predominated in our series, in line with previous reports when studied in regular wards [8,31]. By contrast, critical care studies often have reported a predominance of meningoencephalitic forms [10,20]. This difference likely reflects the wide spectrum of neurological disease and could be resulting from a patient’s delay in seeking healthcare or referral bias, rather than distinct pathophysiological entities. Other neurological manifestations, including stroke and facial diplegia, were consistent with previous case reports [10].

In this series, patients with neuroleptospirosis often lacked leukocytosis and thrombocytopenia. Given that these parameters are indicative of systemic inflammatory response, our findings suggest that forms that present with early neurological involvement, predominantly meningitic, may be associated with a less inflammatory phenotype. Another hypothesis could be that the time to presentation to hospital for a diagnosis with neurological involvement may coincide with the evolution of the disease: normalization of leukocyte counts and platelet recovery. In addition, severe thrombocytopenia constitutes a contraindication to lumbar puncture, which was required for the definition of neuroleptospirosis in this study, and may therefore have introduced a classification bias favoring patients with milder hematological abnormalities.

CSF findings were consistent with previous reports, typically showing a moderate pleocytosis, mild protein elevation, and normal glucose levels [8,31,35]. As previously described, the CSF leukocyte profile evolved over time, with a predominance of neutrophils during the first week, followed by a lymphocytic predominance in the second week, revealing an evolution within inflammatory response [35,36]. Importantly, both profiles remain compatible with neuroleptospirosis [37]. These CSF features closely mimic those of viral meningitis, underscoring the need to systematically consider leptospirosis in the differential diagnosis of aseptic meningitis in endemic settings [7,12].

Among neuroleptospirosis cases, serological diagnoses were more frequent than in non-neurological forms, although PCR remained the primary diagnostic method. This finding is consistent with the biphasic course of leptospirosis, in which neurological involvement preferentially occurs during the immune phase, when blood PCR sensitivity is reduced [1,34]. In line with this, Camous *et al*. reported that neuroleptospirosis accounted for most serological diagnoses, whereas a high proportion of PCR assays were negative [38]. CSF PCR was not systematically performed in our study, due to concerns about its diagnostic performance. Previous studies have reported CSF PCR sensitivities ranging from 60% to 90% [39,40]. Leptospiral bacterial loads can be up to 5-10-fold higher in CSF than in blood, and sometimes CSF PCR may remain positive despite a negative blood PCR [41]. CSF PCR could therefore improve early diagnosis, particularly when serology is still negative or CSF pleocytosis is absent.

Regarding neuroimaging, cerebral infarct has already been reported in leptospirosis, while pachymeningitis has only been described in another spirochetal infection, syphilis [20,42]. While post-lumbar puncture contrast enhancement could account for pachymeningitis, the facial nerve enhancement observed on MRI, consistent with the clinical presentation of facial diplegia, seemed directly related to neuroleptospirosis.

Patients with neuroleptospirosis were significantly younger than those without neurological involvement. This finding is consistent with previous studies describing leptospiral meningitis as predominantly affecting young individuals [12,43]. Age-related differences in immune responses, through their effects on blood-brain barrier permeability, may contribute to differential neurotropism [44]. Alternatively, younger patients presenting with meningeal symptoms may be more likely to undergo lumbar puncture, which could have increased the detection of neuroleptospirosis in this age group.

Two contrasting prognosis profiles of neuroleptospirosis are reported across published series, with isolated meningitic forms generally associated with favorable outcomes, while encephalitic presentations, particularly those accompanied by impaired consciousness, have consistently been linked to poor outcome [31,45]. Studies conducted in intensive care settings have identified neuroleptospirosis as an independent risk factor for mortality, with some deaths directly attributable to severe neurological complications, such as cerebral edema or immune-mediated processes, including acute disseminated encephalomyelitis [19,38]. In contrast, outcomes in our study were consistently favorable. This likely reflects the predominance of isolated meningitic forms, younger patient age, and fewer markers of systemic severity, including thrombocytopenia and hepatorenal dysfunction [46–48]. A survivorship bias cannot be excluded, as the most severe cases may have died before undergoing CSF analysis or neurological investigations.

This study has several limitations. Its retrospective design exposes it to information, selection and classification biases. The exclusion of ambulatory cases, the use of stringent objective inclusion criteria, and the non-systematic use of lumbar puncture may have led to an underestimation of the proportion of neuroleptospirosis. The reliance on serology to define probable leptospirosis may be questioned; however, this limitation was mitigated using a high antibody titer threshold (> 100 IU/L). Microbiological CSF analysis was not systematically performed in all cases of aseptic meningitis. This may have led to underestimation of neurological involvement, particularly in early presentations without pleocytosis or in patients with contraindications to lumbar puncture, thereby favoring the inclusion of patients with milder hematological profiles, notably less severe thrombocytopenia. In addition, two patients were classified as neuroleptospirosis cases based on CSF abnormalities other than pleocytosis (increased CSF lactate or IgG levels), which may be subject to debate. A sensitivity analysis excluding these cases was therefore performed; although it affected the multivariate model, crude odds ratios remained stable. The relatively small sample size limited statistical power and restricted multivariable analyses to an exploratory approach.

Overall, in this endemic setting, neurological involvement accounted for approximately one tenth of hospital-managed leptospirosis cases, most often presenting as meningitis at disease onset. Neuroleptospirosis primarily affected younger patients and was associated with a less inflammatory biological profile, closely mimicking viral meningitis. In contrast to reports from critical care settings, prognosis of neuroleptospirosis was favorable, likely reflecting the predominance of isolated meningitic forms. Distinguishing these milder presentations from severe encephalitic forms may reconcile divergent prognosis data in the literature. Prospective collaborative studies incorporating systematic CSF analysis and microbiological testing are warranted to further characterize the pathophysiology and optimize diagnostic strategies for neuroleptospirosis.

## Data Availability

All data produced in the present work are contained in the manuscript.

## Acknowledgements

The authors would like to thank all the paramedics and physicians from various specialties and from all hospitals throughout French Guiana who were involved in the management of patients included in this study, across the three hospital centers of Cayenne (Centre Hospitalier de Cayenne, CHC), Kourou (Centre Hospitalier de Kourou, CHK), and Saint-Laurent-du-Maroni (Centre Hospitalier de l’Ouest Guyanais, CHOG), as well as in the Remote Prevention and Care Centers (in particular Dr. Mathilde Boutrou and Dr. Celine Michaud). They also acknowledge the biologists responsible for microbiological and molecular diagnostics, particularly Prof. Magalie Pierre-Demar & Dr. Vincent Sainte-Rose in Cayenne, Dr. Angelique Procureur and Dr. Thierry Carage in Kourou, Dr. Jean-François Carod in Saint-Laurent-du-Maroni, the Pasteur Institute of Cayenne (Dr. Magali Dodemont), the Cerba (Dr. Veronique Robert) and Eurofins Biomnis laboratory (Dr. Anne Ovize & Dr. Gerard Perazza). They acknowledge Mathieu Picardeau from the National Reference Center for Leptospirosis for his valuable review, as well as the Center’s collaboration in MAT testing. They are also grateful to all collaborators from the Medical Information Department: Dr. Jean-Michel Cauvin (Cayenne), Dr. Dinh-Vank (Kourou), Dr. Balthazar Ntab (Saint-Laurent-du-Maroni). They express their gratitude to the members of the Department of Research, Innovation and Public Health (DRISP) of Cayenne for their support with regulatory procedures. They are thankful to Dr. Laurent Camous (University Hospital Center of Guadeloupe, French West Indies) for providing access to the detailed neurological data from patients included in the Intensive Care study published in *Critical Care Explorations* in 2024, which were invaluable for the discussion and for comparison of our data with the existing literature. They express their sincere gratitude to all individuals who contributed to data collection or analysis: Dr. Louise Caillard-Humeau, Dr. Deborah Porez, Dr. Geoffrey Grotta, Dr. Gaspard de Moustier, Dr Najibullah Rasouly and Sophie Manuel.

## Supplementary data

**Appendix 1.** Reference panel of the 25 serogroups used for the microscopic agglutination test (MAT) by the National Reference Center of Leptospirosis (Pasteur Institute, Paris)

| Serogroups (25 antigens) |  |  |  |
| --- | --- | --- | --- |
| Australis | Autumnalis | Ballum | Bataviae |
| Canicola | Castellonis | Celledoni | Copenhageni |
| Cynopteri | Djasiman | Grippotyphosa | Hardjo |
| Hebdomadis | Icterohaemorrhagiae | Javanica | Louisiana |
| Mini | Panama | Pomona | Pyregenes |
| Sarmiento | Sejroe | Shermani | Tarassovi |
| Patoc (saprophytic, negative control) |  |  |  |

**Appendix 2.** Cerebrospinal fluid features (n=20) in patients with neuroleptospirosis (n=18)

| Biological parameters of all the cerebrospinal fluid (1<sup>st</sup> or 2<sup>nd</sup>) |  |  |  |
| --- | --- | --- | --- |
| CSF WBC/mm<sup>3</sup> (n=20) | 42 [13-240] | CSF protein, g/l (n=20) | 0.51 [0.32-0.88] |
| PMNs, % (n=13) | 25 [8-78] | CSF glucose, mmol/l (n=20) | 3.15 [3-3.66] |
| Lymphocytes, % (n=13) | 68 [18-89] | CSF IgG, mg/l (n=12) | 86 [38-218] |
| CSF RBC/mm<sup>3</sup> (n=19) | 10 [1-270] | CSF lactate, mmol/l (n=14) | 2.5 [1.7-3.2] |
| CSF leukocyte profile according to the delay of lumbar puncture (1<sup>st</sup> or 2<sup>nd</sup>) |  |  |  |
| D2 | Neutrophilic | D7 | Lymphocytic |
| D4 | Neutrophilic | D8 x 2 | Lymphocytic |
| D6 | Lymphocytic | D10 | Lymphocytic |
| D6 | Mixed | D12 | Lymphocytic |
| D7 | Neutrophilic | D15 | Lymphocytic |
| D7 | Neutrophilic | D17 | Lymphocytic |

Results are presented as medians and interquartile ranges (25<sup>th</sup>-75<sup>th</sup> percentiles), as all variables were continuous. **PMNs**, polymorphonuclear neutrophils; **RBC**, red blood cell count; **WBC**, white blood cell count; **D**, day; **CSF**, cerebrospinal fluid; **IgG**, immunoglobulin G.

**Appendix 3.**
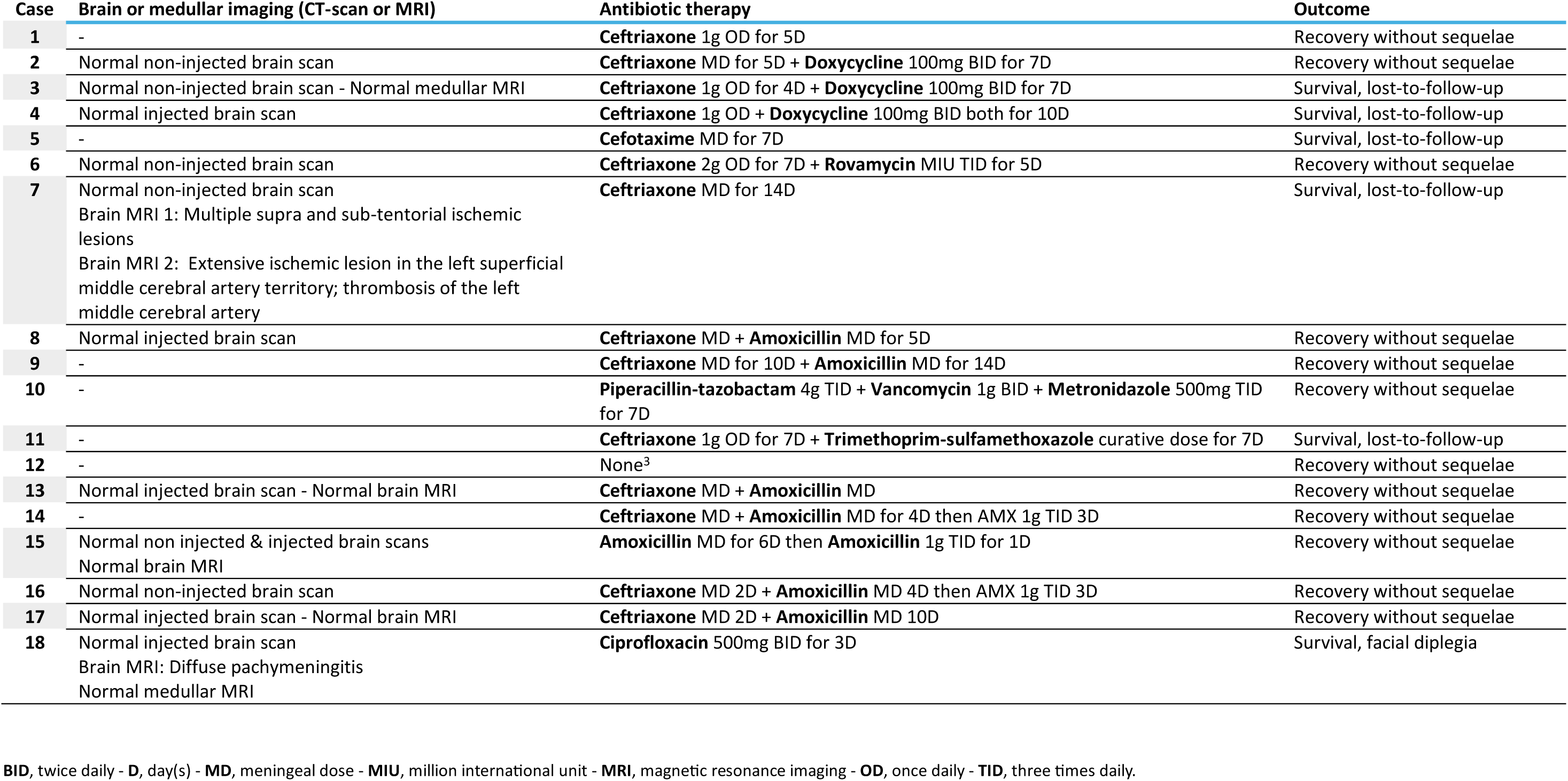
Radiological, and prognostic characteristics of the 18 cases of neuroleptospirosis.

| Case | Brain or medullar imaging (CT-scan or MRI) | Antibiotic therapy | Outcome |
| --- | --- | --- | --- |
| 1 | - | **Ceftriaxone** 1g OD for 5D | Recovery without sequelae |
| 2 | Normal non-injected brain scan | **Ceftriaxone** MD for 5D + **Doxycycline** 100mg BID for 7D | Recovery without sequelae |
| 3 | Normal non-injected brain scan - Normal medullar MRI | **Ceftriaxone** 1g OD for 4D + **Doxycycline** 100mg BID for 7D | Survival, lost-to-follow-up |
| 4 | Normal injected brain scan | **Ceftriaxone** 1g OD + **Doxycycline** 100mg BID both for 10D | Survival, lost-to-follow-up |
| 5 | - | **Cefotaxime** MD for 7D | Survival, lost-to-follow-up |
| 6 | Normal non-injected brain scan | **Ceftriaxone** 2g OD for 7D + **Rovamycin** MIU TID for 5D | Recovery without sequelae |
| 7 | Normal non-injected brain scan<br>Brain MRI 1: Multiple supra and sub-tentorial ischemic lesions<br>Brain MRI 2: Extensive ischemic lesion in the left superficial middle cerebral artery territory; thrombosis of the left middle cerebral artery | **Ceftriaxone** MD for 14D | Survival, lost-to-follow-up |
| 8 | Normal injected brain scan | **Ceftriaxone** MD + **Amoxicillin** MD for 5D | Recovery without sequelae |
| 9 | - | **Ceftriaxone** MD for 10D + **Amoxicillin** MD for 14D | Recovery without sequelae |
| 10 | - | **Piperacillin-tazobactam** 4g TID + **Vancomycin** 1g BID + **Metronidazole** 500mg TID for 7D | Recovery without sequelae |
| 11 | - | **Ceftriaxone** 1g OD for 7D + **Trimethoprim-sulfamethoxazole** curative dose for 7D | Survival, lost-to-follow-up |
| 12 | - | None<sup>3</sup> | Recovery without sequelae |
| 13 | Normal injected brain scan - Normal brain MRI | **Ceftriaxone** MD + **Amoxicillin** MD | Recovery without sequelae |
| 14 | - | **Ceftriaxone** MD + **Amoxicillin** MD for 4D then AMX 1g TID 3D | Recovery without sequelae |
| 15 | Normal non injected & injected brain scans<br>Normal brain MRI | **Amoxicillin** MD for 6D then **Amoxicillin** 1g TID for 1D | Recovery without sequelae |
| 16 | Normal non-injected brain scan | **Ceftriaxone** MD 2D + **Amoxicillin** MD 4D then AMX 1g TID 3D | Recovery without sequelae |
| 17 | Normal injected brain scan - Normal brain MRI | **Ceftriaxone** MD 2D + **Amoxicillin** MD 10D | Recovery without sequelae |
| 18 | Normal injected brain scan<br>Brain MRI: Diffuse pachymeningitis<br>Normal medullar MRI | **Ciprofloxacin** 500mg BID for 3D | Survival, facial diplegia |

**BID**, twice daily - **D**, day(s) - **MD**, meningal dose - **MIU**, million international unit - **MRI**, magnetic resonance imaging - **OD**, once daily - **TID**, three times daily.

**Appendix 4.** Analysis of serogroups identified among 25 positive MAT serologies.

| **Neuroleptospirosis group: 23 positive MAT serologies** |  |  |  |  |  |  |  |  |  |  |
| --- | --- | --- | --- | --- | --- | --- | --- | --- | --- | --- |
| Identified serogroup | Autumnalis | Ballum | Canicola | Copenhageni | Castellonis | Icterohaemorrhagiae | Patoc | Pyrogenes | Sejroe | Serogroup considered responsible |
| P1 | 100 |  | 400 |  |  |  | 400 | 800 |  | Pyrogenes |
| P2 |  |  | 3200 |  | 100 | 200 | 400 | 800 |  | Canicola |
| P3 |  |  | 1600 | 800 | 200 | 800 | 200 |  |  | Canicola |
| P4 |  |  |  |  |  | 3200 | 3200 |  |  | Icterohaemorrhagiae |
| P5 |  |  | 800 |  |  |  |  |  |  | Canicola |
| P6 |  |  | 3200 |  |  |  |  |  |  | Canicola |
| P7 |  |  |  | 3200 |  | 3200 |  |  |  | - |
| P8 |  |  |  |  |  | 800 |  |  |  | Icterohaemorrhagiae |
| P9 |  |  | 3200 |  |  | 3200 | 3200 |  |  | - |
| P10 |  |  |  |  |  | 800 |  |  |  | Icterohaemorrhagiae |
| P11 |  |  | 3200 |  |  |  |  |  |  | Canicola |
| P12 |  |  |  |  |  | 1600 |  |  |  | Icterohaemorrhagiae |
| P13 |  |  |  | 800 |  | 400 | 1600 |  |  | Copenhageni |
| P14 |  |  | 1600 |  |  |  |  |  |  | Canicola |
| P15 |  |  | 3200 |  |  |  |  |  |  | Canicola |
| P16 |  |  |  |  |  | 800 | 800 |  |  | Icterohaemorrhagiae |
| P17 |  |  | 3200 | 3200 |  | 3200 | 3200 |  |  | - |
| P18 |  |  |  | 3200 |  |  |  |  |  | Copenhageni |
| P19 |  |  | 400 | 1600 |  | 400 |  |  | 400 | Copenhageni |
| P20 |  | 800 |  |  |  |  | 200 |  |  | Ballum |
| P21 |  |  |  |  |  | 600 | 600 |  | 800 | Sejroe |
| P22 |  |  | 400 |  |  |  |  |  |  | Canicola |
| P23 |  |  | 3200 | 400 | 400 | 400 | 400 |  |  | Canicola |
| **Neuroleptospirosis group: 2 positive MAT serologies** |  |  |  |  |  |  |  |  |  |  |
| P24 |  |  |  |  |  | 1600 |  |  |  | Icterohaemorrhagiae |
| P25 |  |  | 400 |  |  | 400 |  |  |  | - |

Results are expressed as MAT titers, corresponding to the number of serial dilutions required for agglutination of the serum with the relevant serogroup. **MAT**, microscopic agglutination test - **P**, patient.

## References

1. Levett PN. Leptospirosis. Clin Microbiol Rev. 2001;14(2):296–326. DOI: 10.1128/CMR.14.2.296-326.2001 PMID: 11292640

2. Zenou M, Bourhy P, Abboud P, Picardeau M, Epelboin L, Le Turnier P, et al. Evolution of human leptospirosis in French Guiana, 2016-2022. PLoS Negl Trop Dis. 2025 Oct 13;19(10):e0013620. DOI:10.1016/j.mmifmc.2024.11.005 PMID: 41082585

3. Epelboin L, Bourhy P, Le Turnier P, Demar M, Picardeau M, Djossou F et al. A, et al. Leptospirosis in French Guiana and the Guiana shield: Current knowledge in 2016. Soc Pathol Exot. 2017;110: 165–179. DOI:10.1007/s13149-017-0559-9 PMID: 28478544

4. Turnier P Le, Mosnier E, Schaub R, Picardeau M, Djossou F, Epelboin L et al. Epidemiology of human leptospirosis in French Guiana (2007-2014): A retrospective study. American Journal of Tropical Medicine and Hygiene. 2018;99: 590–596. DOI:10.4269/ajtmh.17-0734 PMID: 30039781

5. Ratet G, Veyrier FJ, Fanton d’Andon M, Picardeau M, Boneca IG, Werts C et al. Live Imaging of Bioluminescent Leptospira interrogans in Mice Reveals Renal Colonization as a Stealth Escape from the Blood Defenses and Antibiotics. PLoS Negl Trop Dis. 2014;8: e3359. DOI:10.1371/journal.pntd.0003359 PMID: 25474719

6. Olie SE, Andersen CØ, van de Beek D, Brouwer MC. Molecular diagnostics in cerebrospinal fluid for the diagnosis of central nervous system infections. Clin Microbiol Rev. 2024;37. DOI:10.1128/cmr.00021-24 PMID: 39404267

7. Alexandre Leite De Souza. Neuroleptospirosis: unexplored & overlooked. Indian J Med Res. 2006 Aug; 124(2):125–8.DOI::10.25259/IJMR_20061242_125 PMID: 17015926

8. Panicker JN, Mammachan R, Jayakumar R V. Primary neuroleptospirosis. 2001 Sep;77(911):589–90. DOI: 10.1136/pmj.77.911.589. PMID: 11524519

9. Wang N, Han YH, Sung JY, Lee W Sen, Ou TY. Atypical leptospirosis: an overlooked cause of aseptic meningitis. BMC Res Notes. 2016 Mar 10;9:154. DOI:10.1186/s13104-016-1964-z PMID: 26964740

10. Mathew T, John’ S, Satishchandra P, Mahadevan A, Chickabasaviah Y. Neuroleptospirosis-Revisited: Experience from a tertiary care neurological centre from south India. Indian J Med Res 124, August 2006, pp 155–162. PMID: 17015929

11. Epelboin L, Abboud P, Abdelmoumen K, Zappa M, Djossou F, Vignier N et al. Overview of infectious and non-infectious diseases in French Guiana in 2022. Med Trop et Sante Int. 2023 2023 Feb 17;3(1):mtsi.v3i1.2023.308. [Article in French] DOI:10.48327/MTSI.V3I1.2023.308 PMID: 37389381

12. Silva HR, Tanajura GM, Tavares-Neto J, Linhares Ad A da C, Vasconcelos PFC, Ko AI et al. [Aseptic meningitis syndrome due to enterovirus and Leptospira sp in children of Salvador, Bahia]. Rev Soc Bras Med Trop. 2002;35: 159–165. DOI:10.1590/S0037-86822002000200006 PMID: 12011925

13. Nabity SA, Ara jo GC, Hagan JE, Reis MG, Ko AI, Ribeiro GS et al. Anicteric Leptospirosis-Associated Meningitis in a Tropical Urban Environment, Brazil. Emerg Infect Dis. 2020;26: 2190–2192. DOI:10.3201/EID2609.191001 PMID: 32818405

14. Tattevin P, Tchamgoue S, Belem A, Benezit F, Pronier C, Revest, M. Aseptic meningitis. Rev Neurol (Paris) 2019 Sep-Oct;175(7-8):475–480. DOI: 10.1016/j.neurol.2019.07.005 PMID: 31375286

15. de Souza AL, Sztajnbok J, Spichler A, Carvalho SM, de Oliveira ACP, Seguro AC. Peripheral nerve palsy in a case of leptospirosis. Trans R Soc Trop Med Hyg. 2006;100: 701–703. DOI:10.1016/J.TRSTMH.2005.10.012 PMID: 16487555

16. Khan SA, Dutta P, Borah J, Topno R, Baishya M, Mahanta J et al. Leptospirosis presenting as acute encephalitis syndrome (AES) in Assam, India. Asian Pac J Trop Dis. 2012;2: 151–153. DOI: 10.1016/S2222-1808(12)60034-6

17. Nguyen L, Chimunda T. Neuro-leptospirosis - A batty diagnostic enigma. IDCases. 2023 Mar 5:32:e01731. DOI:10.1016/J.IDCR.2023.E01731 PMID: 36938340

18. Singh P, Gupta AK, Saggar K, Kaur M. Acute disseminated encephalomyelitis subsequent to Leptospira infection. Ann Trop Med Public Health. 2011;4: 133–135. DOI:10.4103/1755-6783.85771

19. Rathinam SR, Namperumalsamy P. Leptospirosis. Ocul Immunol Inflamm. 1999 Jun;7(2):109–18. DOI:10.1076/ocii.7.2.109.4020 PMID: 10420207

20. Craig SB, Prior SJ, Weier SL, Hall-Mendelin, S& M, David B. David B. et al. Leptospirosis: Messing with Our Minds: A Review of Unusual Neurological and Psychiatric Complexities. Zoonoses: Infections Affecting Humans and Animals. [2^nd^ ed.]. Springer, Cham, Switzerland, pp. 1313–1330. DOI:10.1007/978-3-030-85877-3_34-1

21. Zhu Z, Feng J, Dong Y, Jiang B, Wang X, Li F. Cerebral infarct induced by severe leptospirosis-a case report and literature review. BMC Neurol. 2022 Dec 29;22(1):506. DOI:10.1186/S12883-022-03021-5 PMID: 36581810

22. Centers for Disease Control and Prevention (CDC). Leptospirosis - Yellow Book: Health Information for International Travel. Chapter “Leptospirosis.” Washington DC: CDC; 2026.

23. Human leptospirosis: guidance for diagnosis, surveillance and control. WHO, Geneva. 2003.

24. Epelboin L, Bourhy P, Le Turnier P, Demar M, Picardeau M, Djossou F, et al. La leptospirose humaine en Guyane: etat des connaissances et perspectives. BEH. 2017.

25. Turnier P Le, Mosnier E, Schaub R, Picardeau M, Djossou F, Epelboin L, et al. Epidemiology of human leptospirosis in French Guiana (2007-2014): A retrospective study. Am J Trop Med Hyg. 2018;99: 590–596. DOI:10.4269/ajtmh.17-0734 PMID: 30039781

26. Singer M, Deutschman CS, Seymour C, van dr Poll T, Vincent JL, Augus DC, et al. The Third International Consensus Definitions for Sepsis and Septic Shock (Sepsis-3). JAMA. 2016;315: 801–810. DOI:10.1001/JAMA.2016.0287 PMID: 26903338

27. 27. ARDS Definition Task Force, Ranieri VM, Rubenfeld GD, Fan E, Camporota L, Slutsky AS, et al. Acute respiratory distress syndrome: the Berlin Definition. JAMA. 2012 Jun 20;307(23):2526–33. DOI:10.1001/JAMA.2012.5669 PMID: 22797452

28. Wuthiekanun V, Amornchai P, Langla S, Day NPJ, Limmathurotsakul D, Peacock SJ, et al. Antimicrobial disk susceptibility testing of Leptospira spp. using leptospira vanaporn wuthiekanun (LVW) agar. Am J Trop Med Hyg. 2015;93(2):241–3. DOI:10.4269/ajtmh.15-0180 PMID: 26055750

29. Trombert-Paolantoni S, Thomas P, Hermet F, Clairet V, Litou N, Maury L. Leptospirosis screening: performance of the serion elisa classic leptospira IgM KIT. Pathol Biol (Paris). 2010;58(1):95–9. DOI:10.1016/j.patbio.2009.06.008 PMID: 19892494

30. Dreyfus A, Ruf M-T, Goris M, Poppert S, Mayer-Scholl A, Loosli N, et al. Comparison of the Serion IgM ELISA and microscopic agglutination test for diagnosis of Leptospira spp. infections in sera from different geographical origins and estimation of Leptospira seroprevalence in the Wiwa indigenous population from Colombia. PLoS Negl Trop Dis. 2022;16(6):e0009876. DOI:10.1371/journal.pntd.0009876 PMID: 35666764

31. Gancheva GI, Kostadinova MA, Kostadinova PI. Involvement of central nervous system in leptospirosis. J Biomed Clin Res 2009(Volume 2, Number 2):109-114

32. Silva AF da, da Cruz Franco V, Douine M, Margarete do Socorro Mendonça Gomes 6, Yann Lambert 3, Martha Cecília Suárez-Mutis et al. Brazilian Gold Miners Working Irregularly in French Guiana: Health Status and Risk Determinants. Trop Med Infect Dis. 2024 Dec 31;10(1):12. DOI:10.3390/tropicalmed10010012 PMID: 39852663

33. 33. Coffey JH, Dravin I, Dine QC. Swineherd’s disease (aseptic meningitis) due to Leptospira pomona. J Am Med Assoc. 1951 Nov 3;147(10):949–50. DOI:10.1001/jama.1951.73670270001013 PMID: 14873602

34. Bharti AR, Nally JE, Ricaldi JN, Gotuzzo E, Vinetz JM, Peru-United States Leptospirosis Consortium et al. Leptospirosis: a zoonotic disease of global importance. Lancet Infect Dis. 2003 Dec;3(12):757–71. DOI:10.1016/S1473-3099(03)00830-2 PMID: 14652202

35. Beeson PB, Hankey DD. Leptospiral meningitis. AMA Arch Intern Med. 1952 Apr;89(4):575–83. DOI:10.1001/ARCHINTE.1952.00240040054007 PMID: 14902167

36. Edwards GA. Clinical Studies Clinical Characteristics of Leptospirosis’ Observations Based on a Study qf Twelve Sporadic Cases. Am J Med. 1959 Jul;27(1):4–17. DOI: 10.1016/0002-9343(59)90056-7 PMID: 13661183

37. Silva M V., Camargo ED, Batista L, Vaz AJ, Ferreira AW, Barbosa PRS. Application of anti-leptospira ELISA-lgM for the etiologic elucidation of meningitis. Rev Inst Med Trop Sao Paulo. 1996 Mar-Apr;38(2):153-6. DOI:10.1590/S0036-46651996000200011 PMID: 9071036

38. Camous L, Pommier JD, Tressières B, Carles M, Demoule A, Breurec S, et al. Organ Involvement Related to Death in Critically Ill Patients with Leptospirosis: Unsupervised Analysis in a French West Indies ICU. Crit Care Explor 2024 Jul 8;6(7):e1126. DOI:10.1097/CCE.0000000000001126 PMID: 38980049

39. Ciurariu E, Prodan-Barbulescu C, Mateescu DM, Sorop VB, Susan M, Varga NI et al. Diagnostic Advances in Leptospirosis: A Comparative Analysis of Paraclinical Tests with a Focus on PCR. Microorganisms 2025 Mar 15;13(3):667. DOI: 10.3390/microorganisms13030667. PMID: 40142559

40. Romero EC, Blanco RM, Yasuda PH. Aseptic meningitis caused by Leptospira spp diagnosed by polymerase chain reaction. Mem Inst Oswaldo Cruz. 2010 Dec;105(8):988–92. DOI:10.1590/S0074-02762010000800007 PMID: 21225195

41. Waggoner JJ, Soda EA, Seibert R, Grant P, Pinsky BA. Molecular Detection of Leptospira in Two Returned Travelers: Higher Bacterial Load in Cerebrospinal Fluid Versus Serum or Plasma. Am J Trop Med Hyg. 2015 Aug;93(2):238–40. DOI:10.4269/AJTMH.15-0174 PMID: 26033024

42. Hassin GB, Zeitlin H. Syphilitic cerebral hypertrophic pachymeningitis: Clinicopathologic Studies in a Case. Arch Neurol Psychiatry. 1940;43: 362–371. DOI:10.1001/ARCHNEURPSYC.1940.02280020170015

43. Alston J, Broom J, Doughty C. Leptospirosis in man and animals. 1958

44. Erickson MA, Banks WA. Age-Associated Changes in the Immune System and Blood-Brain Barrier Functions. Int J Mol Sci. 2019 Apr 2;20(7):1632. DOI:10.3390/ijms20071632 PMID: 30986918

45. Haake DA, Levett PN. Leptospirosis in humans. Curr Top Microbiol Immunol. 2015:387:65–97. DOI:10.1007/978-3-662-45059-8_5 PMID: 25388133

46. Li D, Liang H, Yi R, Wang W, Zhang Y, Pan P, et al. Clinical characteristics and prognosis of patient with leptospirosis: A multicenter retrospective analysis in south of China. Front Cell Infect Microbiol. 2022 Oct 17:12:1014530. DOI: 10.3389/fcimb.2022.1014530 PMID: 36325463

47. Hochedez P, Theodose R, Olive C, Cesaire R, Picardeau, Cabie A, et al. Factors associated with severe leptospirosis, Martinique, 2010-2013. Emerg Infect Dis. 2015 Dec;21(12):2221-4. DOI:10.3201/eid2112.141099 PMID: 26583702

48. Guerrier G, Hie P, Gourinat A-C, Goarant C, D’Ortenzio E, Missotte I, et al. Association between Age and Severity to Leptospirosis in Children. PLoS Negl Trop Dis. 2013 Sep 26;7(9):e2436. DOI:10.1371/journal.pntd.0002436 PMID: 24086780

